# Vehicle safety tests, rankings, curb weight, and fatal crash rates: automatic emergency brakes associated with increased death rates

**DOI:** 10.1101/2022.12.08.22283253

**Authors:** Leon S. Robertson

## Abstract

The European New Car Assessment Programme (Euro NCAP), the Insurance Institute for Highway Safety (IIHS), and the U.S. National Highway Traffic Safety Administration (NHTSA) each publish safety ratings of new passenger vehicles based on crash test results and crash avoidance technology soon after they are introduced into the market. IIHS alone singles out vehicles for “Top Safety Pick”. The Institute also periodically lists driver death rates of vehicles during their first few years of use, accompanied by assertions that larger, heavier passenger vehicles are safer. Median death rates by vehicle size in the Institute’s data do not support the assertion. This study examines the association of vehicle weight to the risk of all deaths in fatal crashes where specific makes and models were involved, controlling statistically for the Institute’s vehicle safety ratings as well as NHTSA ratings of full-frontal crash tests and rollover propensity, lane retention warnings, adaptive cruise control, and automatic braking. Increased weight is slightly related to a lower fatal risk in the 2014-2017 models but not the 2018-2019 models. Lane retention warnings and IIHS’s higher ratings of crashworthiness are related to reduced death risk. Adaptive cruise control is not associated with fatal crash risk. Automatic emergency braking (AEB) technology rated “superior” is a criterion for an IIHS “Top Safety Pick” but it is correlated to higher fatal crash involvement of vehicles that have the technology as optional or standard equipment. IIHS tests of AEB systems are conducted only at low speeds. The statistical results are not definitive but suggest that a rating system based on modeling death risk prediction from more detailed data from various tests such as Euro NCAP tests of AEB systems could better inform consumer choice of vehicles.

## INTRODUCTION

Tests of passenger vehicle crashworthiness and certain crash avoidance technology in makes and models of new vehicles are conducted by the Insurance Institute for Highway Safety (IIHS) in the U.S. and scores for each vehicle on four-point scales are posted online (IIHS, no date). The U.S. National Highway Traffic Safety Administration (NHTSA) uses 5 stars to rank crashworthiness based on full frontal and side crash tests and rollover propensity but only lists the availability of some crash avoidance technologies on each make-model (NHTSA, no date). The Euro New Car Assessment Program tests both crashworthiness and crash avoidance technology and assigns percentages to the performance of each (Euro NCAP, no date).

In the past, crash tests and research on vehicle factors related to injury had a major influence on the improved crashworthiness of vehicles. Ratings of the safety features of new vehicles are taken seriously by manufacturers. When their vehicles receive the IIHS “Top Safety Pick” or NHTSA’s 5-star rating, they often use the information in their advertising. But the IIHS ratings and top picks are not comparable across all vehicles; they are awarded within size classes. The webpage that discusses the criteria for top picks says. “These awards identify the best vehicle choices for safety within size categories during a given year. Larger, heavier vehicles generally afford more protection than smaller, lighter ones. Thus, a small car that qualifies for an award might not protect its occupants as well as a bigger vehicle that doesn’t earn the award” (Insurance Institute for Highway Safety. No date). The Top Safety Pick web pages for new models in 2022 included 92 vehicles that did well on crash tests, headlight ratings and “superior” on automatic emergency braking (AEB) tests. When a large majority of vehicles are rated highly, the incentive for improvement is removed.

For almost half a century, the role of vehicle size and weight in road safety has been controversial. In 1974, researchers at IIHS analyzed injuries to unbelted occupants in car-to-car crashes based on the physics of size and weight of the colliding vehicles using data from the University of North Carolina Highway Safety Research Center. In those days seat belt use was about 10-15 percent in the U.S. The physics presented in the report predicts that vehicles with interior space for occupants to decelerate in a crash should have lower occupant death rates. Adding weight to a vehicle increases the energy of vehicles moving at the same speed. Kinetic energy is ½ mass multiplied by velocity squared. Therefore, heavier vehicles should result in increased deaths, particularly so to occupants of the lighter-weight vehicle in multi-vehicle collisions but also with other road users to the extent that braking distances are longer for heavier vehicles. The data on fatal crashes available at the time supported the conclusion that weight relative to the size of vehicles should be minimized and that wide variation in the weight of vehicles in use is detrimental to road safety (O’Neill, Joksch, and Haddon, 1974). While there is no doubt that occupants of lighter-weight vehicles in crashes with heavier vehicles suffer more lethally abrupt deceleration, the problem is at least as much the weight of the heavier vehicle as that of the lighter-weight vehicle. The net effect on the total fatal crash rate depends on whether fewer deaths would have occurred if the vehicles in multiple-vehicle crashes were of more similar weights. Vehicle-to-vehicle crashes produce a minority of fatalities in the U.S.; about 35 percent of vehicles in fatal crashes during 2018 were in multiple-vehicle collisions. Deaths also occur when single vehicles strike fixed objects, roll over and collide with road users unprotected by a shell, mainly pedestrians, pedal cyclists, and motorcyclists.

Another IIHS-supported study of 1440 fatal crashes in Maryland during 1971-1972 found that occupant death rates per registered vehicle were higher in vehicles with shorter distances between the front and rear axles (wheelbase) but the reverse was found for vehicles that struck pedestrians. When all fatal crashes were included, passenger cars with wheelbases less than 106 inches were about 10 percent more often involved than those with wheelbases 121 inches or longer but the rate of pickup trucks was 24 percent more than that of the smaller cars. By far the highest fatal crash involvement per registered vehicle involved motorcycles and tractor-trailer trucks. The authors suggested that future studies of vehicle size should include all road users (Robertson and Baker, 1975). Nevertheless, most of the studies of vehicle size and weight in subsequent decades focused on deaths of occupants, usually drivers only, relative to the size and weight of vehicles in two-vehicle crashes. The weight of road vehicles has consequences other than road deaths. Heavier vehicles that are fueled by gasoline or diesel oil deplete more of those resources and emit more carbon dioxide per mile driven, one of the major contributors to global warming.

The OPEC oil boycott in 1973 exposed the vulnerability of the U.S. economy to fuel shortages as long lines formed at fueling stations that often ran out of product. Congress enacted legislation that set standards for fuel economy for passenger vehicles called Corporate Average Fuel Economy (CAFÉ) standards. As implied by the title, each manufacturer had to achieve an increase in average fuel economy among its products or pay a fine. Since fuel economy is strongly influenced by vehicle weight, manufacturers were under pressure to reduce the weight of their vehicles or achieve increased fuel economy by other means. Originally, the standards called for an average of 27.5 miles per gallon for passenger cars and 20.7 miles per gallon for light trucks and were subsequently incremented dependent on vehicle size. In 1996, Congress prohibited the U.S. Department of Transportation from changing the standards and prohibited governmental research on their effects but in 2001 commissioned the National Academy of Sciences to conduct a review of the effect of the standards. A majority of the committee appointed to study the issue agreed that CAFÉ standards resulted in 1300-2600 road deaths that would not have occurred in 1993 but two members of the committee dissented, saying that the data were not adequate to draw such a conclusion (Committee on the Effectiveness and Impact of Corporate Average Fuel Economy (CAFE) Standards, 2002).

Statistical research on motor vehicle size and weight as predictors of fatalities in crashes is complicated by the correlation of size and weight. When predictors of an outcome such as road deaths are strongly correlated, the coefficients in regression models may be distorted (Selvin, 1991). General Motors researchers produced a study of deaths to drivers of vehicles in use during 1975-1989 in two-vehicle crashes that attempted to distinguish the effects of size and weight. When wheelbases of the colliding vehicles were near equal, the authors said, there was a substantial vehicle weight effect on fatalities but when the weights were similar, the effects of wheelbases were negligible. They concluded that weight is more important than size and derisively claimed that research finding reducing weight to increase fuel economy would not have a major effect on road deaths is “the triumph of zeal over science, or perhaps even common sense’’ (Evans and Crick, 1992). Their graphs, however, showed a wide scatter in the death risk among drivers of vehicles of similar wheelbases but different weights. In his retirement, the senior author revealed in a newspaper opinion piece that he is a climate change denier (Evans, 2010).

A subsequent study of 1999-2000 model year passenger cars, vans, and sports utility vehicles involved in all road deaths during the calendar years 2000-2004 found that driver deaths were lower in association with curb weight but all deaths were higher in association with curb weight. Both driver and all deaths were lower in association with turn distance, an indicator of size less correlated to weight than wheelbase, corrected statistically for performance in crash tests and static stability. Better performance in IIHS and NHTSA crash tests as well as higher static stability was also associated with a lower risk of driver and all deaths (Robertson, 2006). A study of changes in the average and dispersion of vehicle weights in association with CAFÉ standards found that the standards likely reduced road deaths by a few hundred annually as of 2005 (Bento, Gillingham, and Roth, 2017).

The mix of vehicles in use is a function of what manufacturers and retail dealers choose to offer and promote for sale and what vehicle buyers are willing to buy. The average vehicle is in the national fleet for substantially more than a decade. Whether buyers are influenced much, if any, by safety ratings or vehicle weight among the myriad characteristics of vehicles in the marketplace is open to question. A 1989 survey of U.S. new vehicle buyers indicated a variety of factors they say influenced the purchase. Weight was not included as a choice in the questions asked but fuel efficiency, a strongly inverse correlate of weight at that time, was deemed desirable. So were size and safety (Tay, 1998). A study of vehicle sales before and after IIHS issued new ratings suggested that the ratings affected sales (Cicchino, 2015).

The most astonishing trend in vehicle mix in the U.S. is the increased market share of sport utility vehicles (SUVs) and pickup trucks. In 1970, only 17 percent of registered vehicles were “light trucks”. The registrations, including SUVs, increased to 28.8 percent in 1990, 39.3 percent in 2000, 45.5 percent in 2010, and 58.5 percent in 2020 (U.S. Department of Transportation, annual). Figure 1 indicates that SUVs substantially replaced passenger cars while pickup truck registrations grew as well. In the 21^st^ century, new pickup trucks are larger and heavier than their predecessors for the most part while SUVs vary widely in size and weight. Based on an analysis of vehicle and driver characteristics in severe injury crashes around the turn of the century, one study concluded that if all cars of the same weight as SUVs and pickup trucks had been the latter vehicles, occupant deaths would have been 26 percent and 64 percent more frequent respectively (Wang and Kockleman, 2005). Another characterized the increase in sales of heavier SUVs and pickup trucks as an “arms race” (White, 2004). A study of pedestrian deaths per number of pedestrians struck found that pickup trucks, SUVs, and large vans were more deadly (Lefler and Gabler, 2004). One study estimated that more than 8000 pedestrian deaths during 2000-2019 were attributable to the growth in SUV and pickup trucks (Tyndall, 2021). Not only weight but incompatibility in the engagement of energy absorption structure are correlated to driver death rates in vehicle-to-vehicle collisions (Monfort and Nolan, 2019).

**Figure 1.**
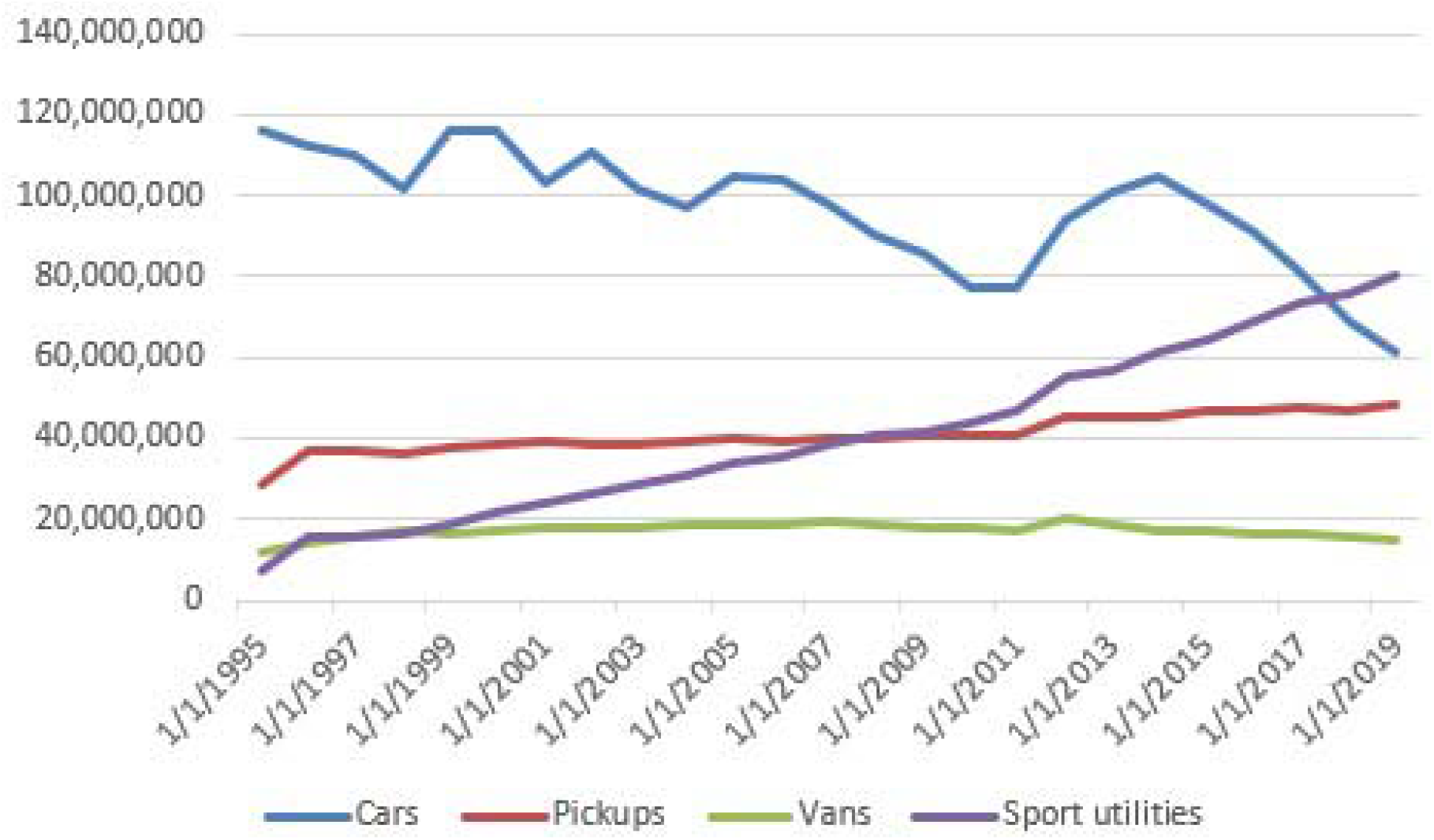
U.S. registered vehicles by type, 1995-2019 Sources: https://fred.stlouisfed.org/series/USASACRQISMEI and https://www.bts.gov/browse-statistical-products-and-data/national-transportation-statistics/number-us-truck

For the past three decades, IIHS periodically published in its newsletter, Status Report, driver death rates by make and model of recently sold passenger vehicles. In a commentary accompanying each publication, vehicle size and weight were emphasized as major factors in the differences in driver death rates. The Institute’s classification of vehicles as “small”, “midsize”, “large”, and “very large” is based on a combination of weight and length times width that the Institute calls “shadow”. There is a large variation in weight within categories. For example, a passenger car classified as “small” can be up to 4000 lbs. in weight but a car classed as “midsized” can be as little as 2500 lbs. in weight (Highway Loss Data Institute, 2020). Since size and weight are highly correlated, the variation of weight within the size classes is not that large in recently manufactured vehicles.

The most recent IIHS publication of variation of driver death rates in these size classes as well as separate ones for SUVs and pickup trucks and “luxury vehicles” included 2014-2017 models in use during 2015-2018. The headline read “Driver death rates remain high among small cars” (Insurance Institute for Highway Safety, 2020). The data in the table, however, showed a huge variation in driver death rates and little relation between the rates and vehicle size. In the “small” passenger car category driver deaths varied from zero to 98 per hundred registered vehicle years with a median of 45. The median for “large” cars was higher, 50, and a little less, 41, for “midsize” passenger cars. A similar pattern occurred among SUVs – the median driver death rates of “small”, “mid-sized” and “large” SUVs were 29, 16, and 25.5 respectively. Such a pattern does not support the “small is bad” rhetoric. Only driver deaths are included under the assumption that the inclusion of all occupants could cause variation in rates because some vehicles may systematically carry more occupants than others. The potential effects of size and weight on the deaths of other road users were ignored. Previous research on the association of vehicle safety ratings and injury rates produced mixed results regarding the predictability of injuries from ratings but none included vehicle weight or recent crash avoidance technology (e.g., Harless and Hoffer, 2007; Lidbe, et al, 2020; Phillips, et al. 2021; Segui-Gomez and Lopes-Valdez, 2007).

The purpose of this paper is to report analyses of vehicle weight, test-based safety ratings, and available crash avoidance technologies as fatal crash risk factors. The results suggest a better means to inform consumers about the relative importance of vehicle factors in road deaths.

## METHODS

Data on fatal crashes in selected calendar years by make and model of vehicles were downloaded from the Fatal Analysis Reporting System (NHTSA File Downloads, 1975-2020). Beginning in 1975, the files contain annual data on crash sites, vehicles, drivers, and others involved in fatal crashes where one or more persons involved died within 30 days of the incident. NHTSA included the curb weight of cars in 1975 and SUVs, and pickups in the files in 2012 but dropped it thereafter. The weight distributions of cars weighing less than 10,000 lbs. in 1978 and 2012 fatal crashes were compared to see if the average and variability of the weight distribution of passenger cars in fatal crashes declined or expanded in the decades after fuel economy standards were adopted.

In an attempt to account for the large variation in death rates by make and model, vehicle curb weight, NHTSA ratings of crashworthiness and rollover propensity, and IIHS ratings of crash test results, headlight performance, and automatic emergency brake (AEB) technology as options were examined for their value in predicting road deaths. Since the Fatal Analysis Reporting System codes by vehicle name do not include some of the distinctions that IIHS uses, where IIHS listed two or more versions of the same named vehicle to distinguish 2-wheel vs. 4-wheel drive and other distinctions, the counts of registered years were added together. Where one or more distinctions of the same make/model were excluded from IIHS data, the make/model was excluded from the analysis of 2014-2017 model year vehicles. Seventy make-models were included.

An attempt to use Poisson regression to assess the relative predictability of crashworthiness and crash avoidance technology was unsuccessful as the model failed to converge. Statistical purists argue that Poisson regression should always be used for the analysis this type of data but logistic regression produces highly similar results to Poisson regression (e.g., Mittlbock and Heinzl, 2001). Therefore, logistic regression was used to assess the risks of deaths by make and model of 2014-2017 models in 2015-2018 associated with the predictor variables. The form of the equation is:

Road deaths/years used (where a given make/model was involved) = B_1_ (curb weight) +

B_2_ (small overlap frontal crash test) +

B_3_ (moderate frontal overlap crash test) +

B_4_ (side crash test) +

B_5_ (median headlight rating)

B_6_ (“Superior” AEB standard or optional) +

B_7_ (“Advanced” AEB standard or optional) +

B_8_(“Basic” frontal crash warning signal standard or optional)

B_9_(1 if pickup truck, else 0)

B_10_(1 if SUV, else 0)

To assess the reliability of the estimates, a second equation was applied to deaths among 2018-2019 models in their first two years of use. The adoption of AEB as standard equipment on 35 vehicles in those model years allowed the assessment of whether a correlation of AEB with increased risk in 2014-2017 models was an artifact of its previous optional status. Front and rollover ratings from NHTSA as well as lane departure warnings and adaptive cruise control as optional or standard equipment were also added to the second equation.

IIHS crash test and headlights are rated on a 4-point scale with the highest (“good”) value indicating safer. Results of roof strength were rated “good” on almost all the vehicles so it was excluded. Side crash tests by NHTSA were excluded for the same reason. When more than one headlight rating was included depending on trim, the median was used in the analysis. The frontal crash prevention technology labels, “superior”, “advanced” and “basic”, are based on IIHS low-speed tests of the initiation and performance of automatic braking or warning. These systems are supposed to detect situations in which braking is needed to avoid or reduce the severity of collisions. Essentially, the “basic” system warns drivers to apply brakes, and the designations “superior” and “advanced” are qualitative categories based on how the systems react as they approach stationary targets at varying low speeds on a test track (Insurance Institute for Highway Safety, no date). When these technologies were optional in most of the 2014-2017 models they were indicated as 1 if available and 0 if not. The few that were standard were also assigned 1. If the option changed from one year to the next, the variable was apportioned in fractions of 1. The percentage of sold vehicles equipped with the technology varied substantially among manufacturers (National Highway Traffic Safety Administration, 2019) but no data on the number sold by make and model were found.

In the analysis of 2018-2019 vehicles in their first two years of use, new vehicle sales by month were copied and pasted from goodcarbadcar.net or manufacturer websites. The number sold in a given month was multiplied by the months they were in use during two years. The sum of the months was divided by 12 to get the total years of use for each model year of each make-model. Since vehicle scrappage is minimal during the first few years of use, this method is likely more accurate than the once-a-year count of registered vehicles. Only vehicles with more than 100,000 years of use were included in the regressions. The presence or absence of lane departure technology and adaptive cruise control on a given make/model was obtained by searching manufacturer and dealer websites. Since it was not always possible to find whether these were standard or optional equipment, they were assigned 1 if available and 0 if not.

## RESULTS

The Fatal Analysis Reporting System does not contain curb weights of the vehicles in fatal crashes after 2012. Figure 2 shows the distribution of curb weights of passenger cars in fatal crashes in 1978 and Figure 3 shows the distribution in 2012.

**Figure 2.**
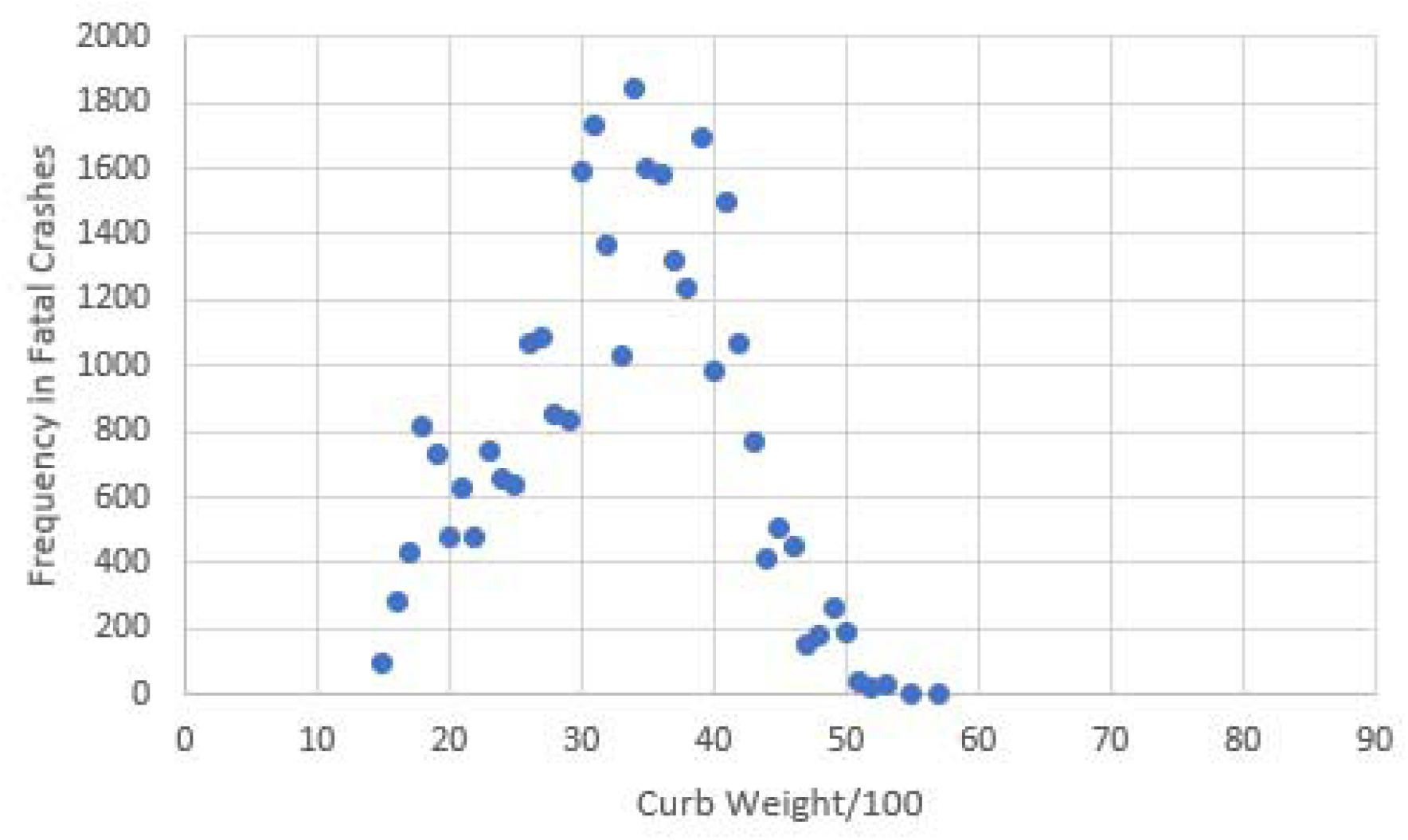
Frequency of passenger cars in fatal crashes by curb weight, United States 1978.

**Figure 3.**
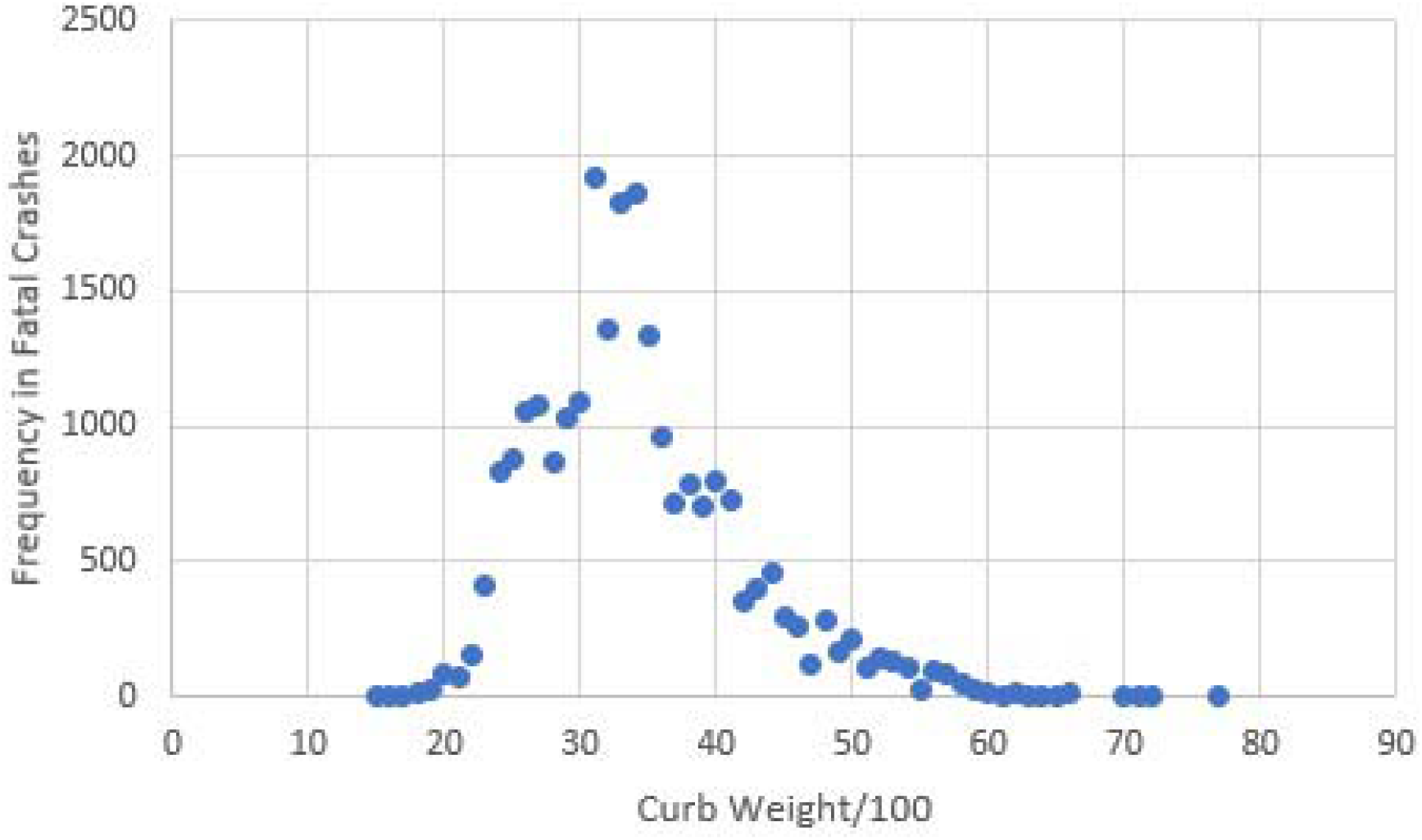
Frequency of passenger cars in fatal crashes by curb weight, United States 2012.

The median curb weights in 1978 and 2012 were similar, about 3300 lbs. but the distribution was wider in 2012. Vehicles such as the original Volkswagen Beetle, Honda Civic, and Chevrolet Chevette weighing less than 2000 lbs. that were popular in the 1960s and 1970s were seldom in use by 2012. The modern Beetle and Civic weigh about 2712 and 2877 lbs. respectively. No passenger cars weighing more than 5800 lbs. were involved in 1978 but several were involved in 2012. The curb weight of pickup trucks and SUVs was not included in the 1978 FARS file so they were excluded from both figures to keep the distributions comparable. Pickup trucks that became more popular over time increased in weight. For example, versions of the 1978 Chevrolet C/K pickups had curb weights from 3700-4000 lbs. but the 2012 versions were about 1500 lbs. heavier. Despite the reduced sales of cars weighing less than 2000 lbs., the dispersion in weights increased substantially as the sales of SUVs and pickup trucks increased.

The percent of vehicle occupants that died in 2-vehicle fatal collisions correlated to differences in curb weights in 2012 are displayed in Figure 4. The wider the weight discrepancy, the more likely that a driver or passenger in the lighter vehicle will be killed. The percentage of occupants in both vehicles that were killed is slightly higher when the vehicles are closer in weight. Figure 5 indicates that the number of occupants per vehicle is not systematically different in the lighter and heavier vehicles in these collisions.

**Figure 4.**
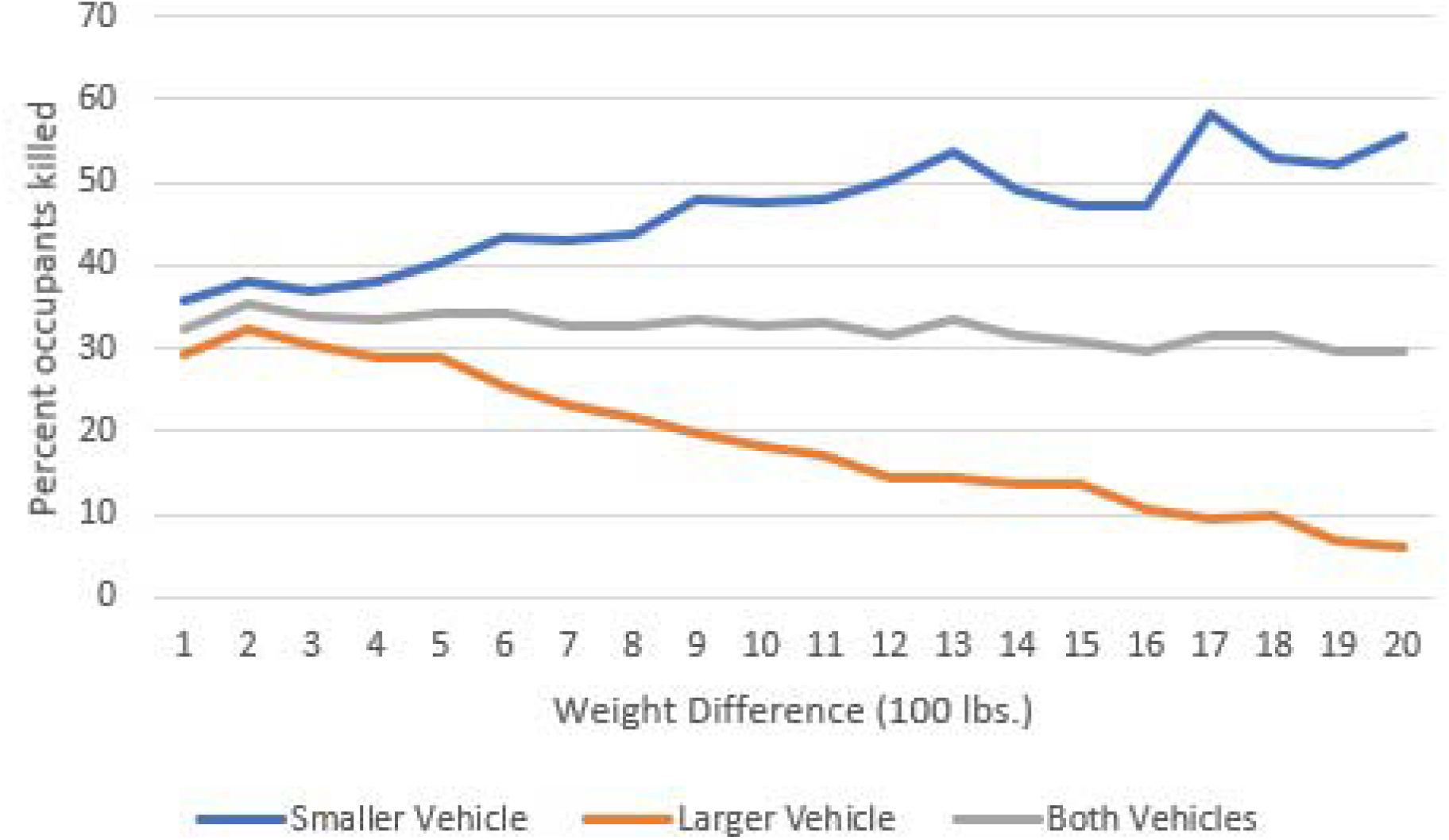
Percent occupants killed in vehicle-to-vehicle collisions by weight difference, 2012

**Figure 5.**
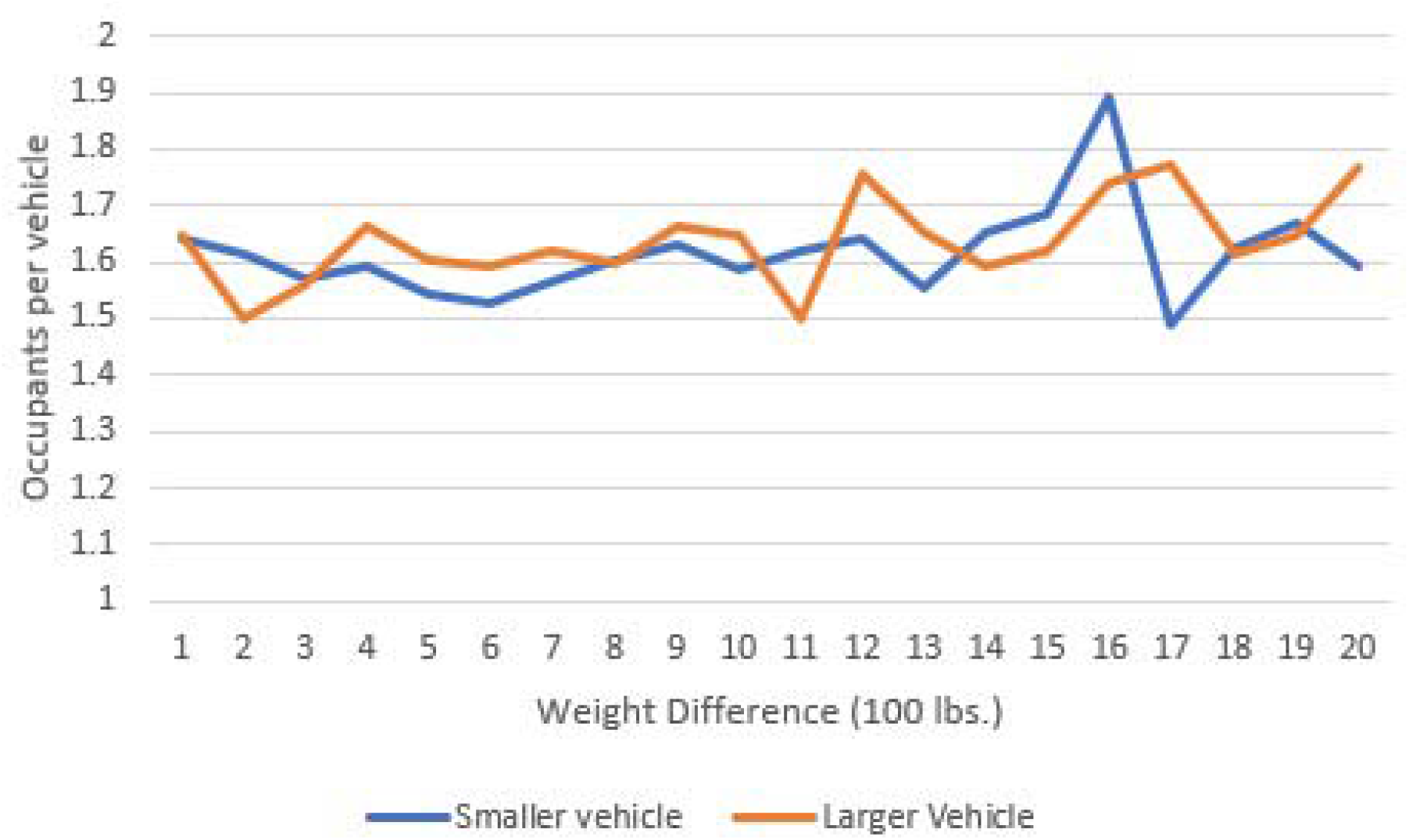
The number of occupants per vehicle in 2-vehicle fatal crashes by the difference in weight between the vehicles.

Driver death rates per million years of use by the curb weight of 2014-2017 vehicles in 2015-2018 are displayed in Figure 6. The scatter is remarkable. A graph (not shown) of driver deaths per million years used by square footage (length x width that IIHS calls “shadow”) as an indicator of size indicates a similar pattern. Square footage and curb weight are so strongly correlated (R^2^= 0.80) that it is not possible to estimate the effect of each independent of the other but neither is related strongly to driver death rates. When all deaths in which vehicles of a given weight are involved (Figure 7), the scatter is similar to that of driver death rates. Vehicle curb weight alone is an unreliable predictor of the driver or total death rates.

**Figure 6.**
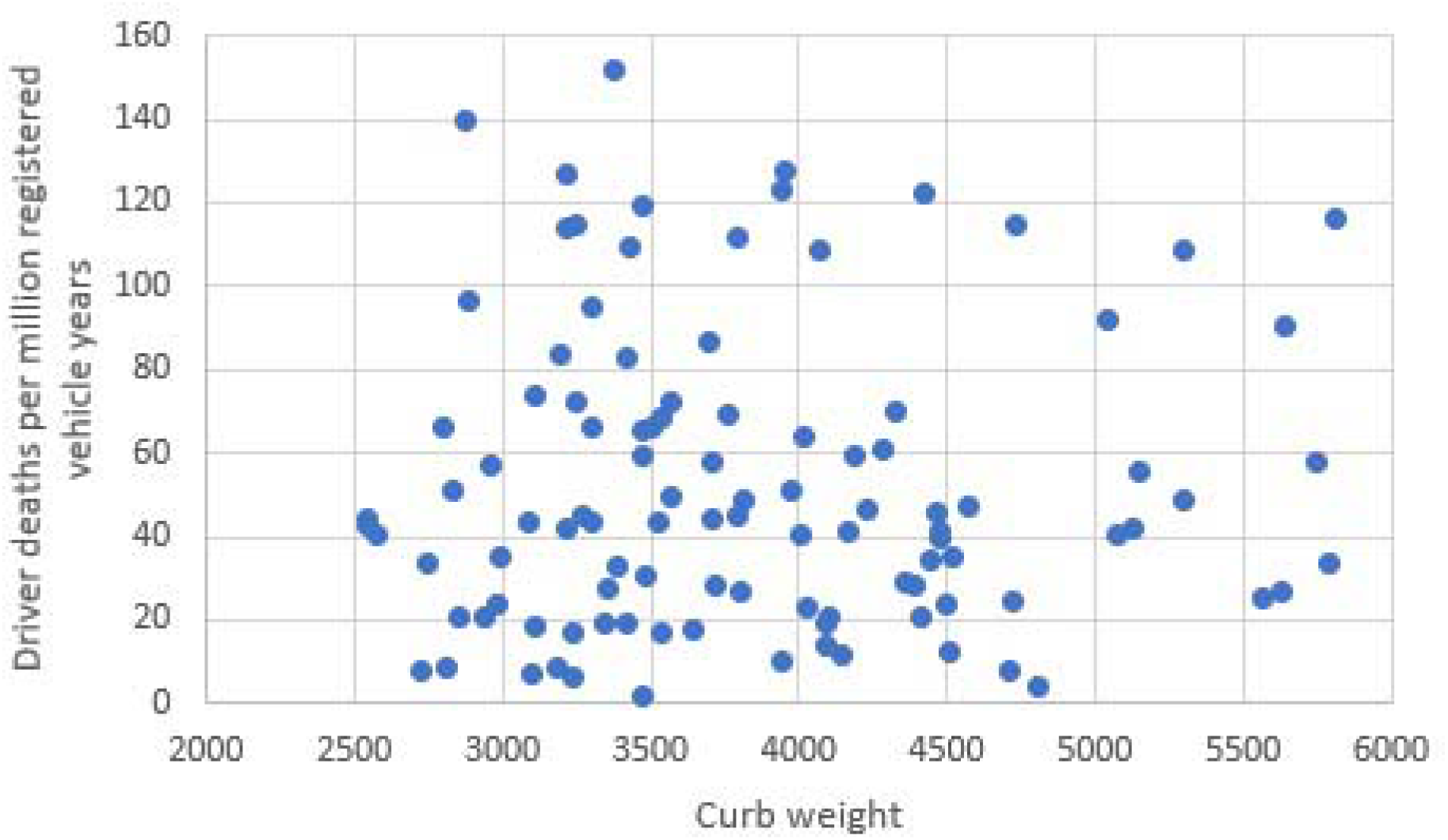
Driver deaths per million registered passenger vehicle years of use by curb weight – 2014-2017 models in 2015-2018.

**Figure 7.**
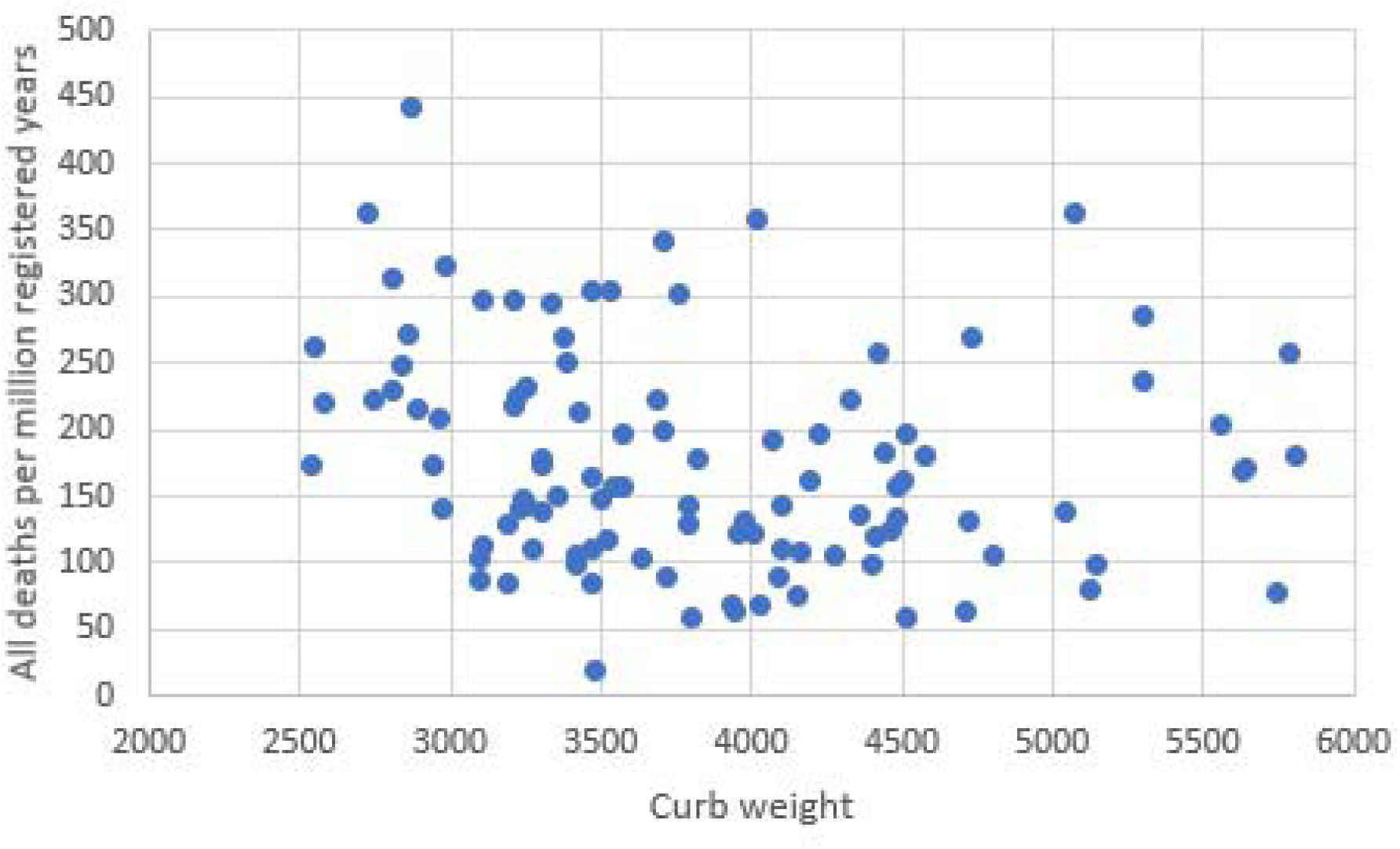
Involvement of passenger vehicles in all deaths per million registered vehicle years by curb weight – 2014-2017 models in 2015-2018

The odds ratios and 95 percent confidence intervals of curb weight and the other predictor variables are presented in Table 1. Corrected statistically for the other predictors, higher weights were slightly correlated to lower fatality risk for the 2014-2017 vehicles but the confidence intervals overlap 1 for the 2018-2019 vehicles. The heaviest vehicles - pickup trucks - have a higher fatal crash involvement risk relative to cars, SUVs, and vans in 2014-2017 but the association was not significant for the 2018-2019 vehicles. SUVs were associated with lower risk in both samples. The IIHS crashworthiness ratings were predictive of lower risk in both samples but the side test was not significant for the 2018-2019 vehicles. NHTSA frontal crash tests and rollover ratings did not add significantly to the predictive value of the equation. Headlight ratings were related to lower risk in 2014-2017 vehicles but not enough to achieve statistical significance in 2018-2019 models. Lane departure warnings were related to lower fatality risk. The fatal crash risk was higher in vehicles with optional “superior” AEB systems in both samples and AEB as standard equipment was predictive of higher risk in 2018-2019 vehicles. “Advanced” AEB was associated with a lower risk among the 2014-2017 vehicles but a higher risk among 2018-2019 vehicles. Adaptive cruise control and “basic” warning of the need to brake added no predictive value. Most of the correlations among the predictor variables were very low. The highest R^2^ was 0.25 between curb weight and pickup trucks. The regression estimates are not likely affected by collinearity among the predictor variables.

**Table 1.** Odds ratios and 95% confidence intervals of predictors of U.S. passenger vehicle fatal crash involvement

| Predictor Variable | 2014-2017 model vehicles during 2015-2018 | 2018-2019 model vehicles during their first 2 years of use |
| --- | --- | --- |
| Curb Weight (100 lbs.) | 0.983 (0.978, 0.988) | 0.995 (0.987, 1.002) |
| IIHS small overlap front crash | 0.895 (0.874, 0.916) | 0.904 (0.852, 0.960) |
| IIHS moderate overlap front crash | 0.974 (0.950, 0.998) | 0.850 (0.755, 0.956) |
| IIHS side crash | 0.951 (0.930, 0.973) | 0.970 (0.928, 1.015) |
| IIHS headlights | 0.900 (0.882, 0.918) | 1.012 (0.958, 1.069) |
| NHTSA full front crash |  | 0.995 (0.912, 1.085) |
| NHTSA rollover |  | 1.021 (0.966, 1.080) |
| Lane departure warning (mostly standard) |  | 0.858 (0.801, 0.919) |
| Adaptive cruise control (mostly standard) |  | 0.982 (0.850, 1.135) |
| IIHS “Basic” Frontal Crash Warning | 1.056 (0.998, 1.116) | 0.860 (0.729, 1.017) |
| AEB Standard |  | 1.336 (1.121, 1.593) |
| IIHS “Superior” AEB (mostly optional) | 1.153 (1.085, 1.225) | 1.338 (1.203, 1.601) |
| IIHS “Advanced” AEB (mostly optional) | 0.816 (0.752, 0.885) | 1.785 (1.351, 2.357) |
| SUVs | 0.710 (0.679, 0.743) | 0.638 (0.576, 0.707) |
| Pickup trucks | 1.636 (1.494, 1.791) | 1.102 (0.935, 1.297) |

## DISCUSSION

The IIHS inclusion of AEB systems as criteria for increased safety is not supported by these results. The IIHS tests of AEB technology are conducted at 12 and 25 miles per hour. In 2014, The National Highway Traffic Administration (NHTSA) performed AEB tests on four 2014-2015 model vehicles up to an approach speed of about 30 miles per hour. Of eight tests on each vehicle, only one met the criteria for reliable repeated performance (Forkenbrock and Snyder, 2015). Nevertheless, NHTSA and IIHS persuaded some 20 vehicle manufacturers to agree to provide AEB as standard equipment by 2025. Most did so earlier and almost all 2022 models have the system. A 2013 proposal to add AEB testing to NHTSA’s New Car Assessment Program was not implemented but remains under consideration (Homedy, 2022). Manufacturers are likely to design their vehicles to get good ratings on NHTSA and IIHS tests. In 2022, the American Automobile Association reported tests of several AEB systems at 30 and 40 miles per hour. At the higher speed, the system failed to prevent a front-to-rear collision with a stationary plastic vehicle mockup in 14 of 20 test runs but reduced the speed of the crash by 62 percent on average. The system failed to prevent collisions at T intersections or when the vehicle was turning left across the path of an oncoming vehicle (AAA, 2022).

If AEB tests are not predictive of risk in the range of conditions that increase crash severity, they are not likely to reduce deaths, and the potential for harm cannot be ruled out. Interviews with early AEB adopters indicated that most leave the system turned on, believe that it had prevented crashes, and intended to have it when they purchased their next vehicle. The authors of that study noted that too much dependence on AEB by drivers or driver reactions to it should be more thoroughly researched (Cicchino and McCartt, 2015). A joint IIHS-MIT study of driver behavior during four weeks in more automated vehicles found increased use of cell phones and willingness to have hands off the wheel as they became familiar with the systems (Regan et al., 2021).

The lower risk of fatal crashes associated with lane departure warnings and increased risk associated with automated braking suggest the hypothesis that warnings increase driver alertness and attention but that automation reduces them. Based on regression analysis of numerous studies of insurance claims data, the Highway Loss Data Institute (HILDI) concluded the opposite. Several factors were included in the HILDI regression model in addition to crash avoidance technologies but neither curb weight nor IIHS and NHTSA ratings of crashworthiness were listed among them. There are large differences in the association of crash avoidance technology and insurance claims by type of coverage (HILDI, 2020). While the reasons for the differences in results using insurance claims data and the death data are not obvious, making an insurance claim for minor injury or property damage is discretionary while reporting a death is not. Litigation of liability claims can take years and insurance claims can be fraudulent.

Other researchers used a variety of comparisons in statistical studies that found AEB efficacious. Some of the earliest studies used “induced exposure” -- a comparison of crashes in which the technology should reduce crashes relative to other crash involvement of the same vehicles (Rizzi, Kullgren, and Tingvall, 2014; Fildes et al., 2015). If the technology increases the risk of those other crashes, inference of the effectiveness of the technology is compromised at best and misleading at worst. A study of rear-end crashes exclusively found that AEB was less likely to prevent a collision when either vehicle was turning rather than going straight and on roads with 70+ mph speed limits or on roads slickened by ice or snow. The system was less effective when the struck vehicle was not a passenger vehicle. Because most rear-end crashes are on dry roads with lower than 70 mph speed limits when both vehicles are going straight, the overall rear-end crash rate of vehicles with AEB was lower (Cicchino and Zuby, 2019). A study of an AEB system to detect pedestrians and slow or stop the vehicle automatically found that it was not associated with a reduction in nighttime pedestrian injuries (Cicchino, 2022) so IIHS recommends its use only during daytime. IIHS researchers have also suggested the use of driver monitoring technology that warns when inattention or lack of hand on the steering wheel occurs (Mueller, Reagan, and Cicchino, 2021). But what if AEB increases the risk of various types of crashes for reasons other than driver disengagement? An increase in U.S. Road deaths in 2020 was correlated mainly to increased sales of pickup trucks, lower fuel prices, and warming temperatures (Robertson, 2022) but increased numbers of vehicles with AEB systems may have contributed as well. In some of the vehicles with AEB, the driver has the choice of setting the system at warning only or turning it off. In others, the system is reset to “on” each time the vehicle is started so the driver must actively turn it off to prevent its use. A case-control study of vehicles in all fatal crashes compared to those at the same sites, same time of day, and day of the week can better discern the effect of technology as well as behavior such as speeding (Robertson and Maloney, 1997). Such studies of the types and usage of AEB technology are sorely needed.

The results support the conclusion that promoting the use of heavier vehicles is not a means to reduce road deaths. With or without correction for crashworthiness and crash avoidance technology, there is little or no correlation between higher weight and reduced risk of total fatalities but the disparity in weights increases the risk to occupants in lighter vehicles in multi-vehicle crashes. Contrary to expectations when the original fuel economy regulations were proposed, vehicle weights of vehicles in use increased over time as manufacturers found ways to increase fuel economy other than reducing weight and marketed heavier cars, trucks, and SUVs. A 2009 U.S. government proposal to increase the averages in fuel economy was met with the same old argument that it would increase road deaths but several analysts found otherwise. Given electric and hybrid gas-electric technology, fuel economy will be less dependent on vehicle weight as vehicles with such technology replace older vehicles without it (Robertson, 2018).

There is no way to know how much the IIHS emphasis on size and weight influences manufacturer or consumer choices as opposed to other factors that influence them. On its “Vehicle size and weight” webpage, the Institute says that larger-sized and weighted vehicles are safer but acknowledges that the differences among driver death rates correlated to square footage narrowed in each iteration of the Institute’s listing of such death rates by size (Insurance Institute for Highway Safety, no date). Since the vehicles in IIHS data tables have been in use for 2 or more years by the time the data are published, they may not have much influence on new vehicle purchases but the Institute’s message that big heavy vehicles are safer vehicles is reported in car buff magazines (e.g., Brady and Lugo, 2012) and other media outlets frequently, except for a few dissenters who notice that there is huge variability in driver death rates in the IIHS lists (e.g., Bestride staff, no date).

Current safety ratings of new vehicles on the IIHS website indicate whether AEB is standard equipment, optional or missing but the Institute does not discuss the relative extent of risk reduction provided by these or the crash test and headlight ratings on each page devoted to specific makes and models. Lane departure warning systems related to lower risk are not included in the ratings. A consumer has no way to evaluate which ratings are the more important.

To achieve accountability by manufacturers and better inform consumers, more empirically derived and discriminating criteria must be employed in vehicle safety ratings. The regression analysis of IIHS and NHTSA test results in this paper is not definitive because some of the differences among vehicles are embedded in aggregated data where the technology was optional and the ratings on all of the variables except weight are qualitative summaries of quantitative data. But the analysis illustrates a means by which IIHS tests could be used to develop a more valid assessment of road death risk among vehicles. A better model would employ quantitative data from test dummy instrumentation and control for weight or size by retaining it in the model rather than showing separate results by arbitrary size categories. AEB tests should be expanded to include speeds at which more severe injuries and deaths occur and should be repeated numerous times each to gauge system reliability. To the extent possible, a model that scores each vehicle based on the probability of a reduction in all fatalities related to individual technologies could influence manufacturer and consumer choices to reduce road deaths. The resulting predictions of death risk for each make and model could be summarized in a single score expressed as a percentile. Anyone who has attended school should understand a grading system that varies from zero to 100. If the “Top Safety Pick” distinction is retained, it could be based on the vehicles that score above a selected percentile. The European vehicle testing agency rates vehicle components on percentage scales but does not combine them into an overall risk score. The most recent ratings on AEB varied wisely, from 14 to 95 percent (Euro NCAP, no date) based on a much more detailed testing protocol (Euro NCAP, 2019) than that used by IIHS. Whether the higher-scoring AEB systems reduce death and injury could be investigated by a case-control study in Europe.

## Data Availability

The data are from public downloadable file referenced in the manuscript.

## REFERENCES

AAA (2022). Evaluation of Emergency Braking Systems. https://newsroom.aaa.com/wp-content/uploads/2022/09/Research-Report-2022-AEB-Evaluation-FINAL-9-26-22-1.pdf

Bento, A., Gillingham K., Roth K. (2017). The effect of fuel economy standards on vehicle weight dispersion and accident fatalities. Bureau Of Economic Research. 2017. https://www.nber.org/system/files/working_papers/w23340/w23340.pdf

Bestride staff. (No date). MYTH BUSTED – Larger, heavier vehicles are not necessarily safer. Bestride.com. https://bestride.com/news/safety-and-recalls/myth-busted-larger-heavier-vehicles-are-not-necessarily-safer

Boyd, P.L. (No date). NHTSA’s NCAP rollover resistance rating system. Washington DC: National Highway Traffic Safety Administration. https://www-esv.nhtsa.dot.gov/Proceedings/19/05-0450-O.pdf

Brady, D. and Lugo R. (2012). Are bigger cars safer than smaller cars? What about SUVs and trucks? Motor Trend. December 22. https://www.motortrend.com/news/are-bigger-cars-safer/

Cicchino, J.B. (2015). Consumer response to vehicle safety ratings. Proceedings of the 24th International Technical Conference on the Enhanced Safety of Vehicles. Washington, DC: National Highway Traffic Safety Administration. https://www-esv.nhtsa.dot.gov/Proceedings/24/files/24ESV-000069.PDF

Cicchino, J.B. (2022). Effects of automatic emergency braking systems on pedestrian crash risk. Accident Analysis and Prevention. 172:106686. Effects of automatic emergency braking systems on pedestrian crash risk - ScienceDirect

Cicchino, J.B. and McCartt, A.T. (2015). Experiences of model year 2011 Dodge and Jeep owners with collision avoidance and related technologies. Traffic Injury Prevention. 16:298–303. https://scholar.google.com/scholar?hl=en&as_sdt=0%2C3&q=Characteristics+of+rear-end+crashes+involving+passenger+vehicles+with+automatic+emergency+braking&btnG= Experi ences of Model Year 2011 Dodge and Jeep Owners With Collision Avoidance and Related Technologies: Traffic Injury Prevention: Vol 16, No 3 (tandfonline.com)

Cicchino J.B. and Zuby D.S. (2019). Characteristics of rear-end crashes involving passenger vehicles with automatic emergency braking. Traffic Injury Prevention. 20:S1,S12-S18. https://scholar.google.com/scholar?hl=en&as_sdt=0%2C3&q=Characteristics+of+rear-end+crashes+involving+passenger+vehicles+with+automatic+emergency+braking&btnG=

Committee on the Effectiveness and Impact of Corporate Average Fuel Economy (CAFE) Standards. (2002). Effectiveness and Impact of Corporate Average Fuel Economy (CAFE) Standards. Washington, DC: National Academy Press.

Euro NCAP. (No date). Latest safety ratings. Euro NCAP | Latest Safety Ratings. Accessed

Euro NCAP. (2019). Test protocol – AEB Car-to-Car Systems. https://cdn.euroncap.com/media/56143/euro-ncap-aeb-c2c-test-protocol-v302.pdf

Evans L. (2010). Has global warming got you snowed in? The Washington Times. https://www.almendron.com/tribuna/has-global-warming-got-you-snowed-in/

Evans L. and Crick M.C. (1992). Car size or car mass: Which has greater influence on fatality risk? American Journal of Public Health. 82:1105–1112. https://ajph.aphapublications.org/doi/pdf/10.2105/AJPH.82.8.1105

Fildes B. et al. (2015). Effectiveness of low speed autonomous emergency braking in real-world rear-end crashes. Accident Analysis and Prevention. 81:24–29. https://www.sciencedirect.com/science/article/abs/pii/S0001457515001116

Forkenbrock G.J. and Snyder A.S. (2015). NHTSA’s 2014 automatic emergency braking test track evaluations. (Report No. DOT HS 812 166). Washington, DC: National Highway Traffic Safety Administration. https://www.nhtsa.gov/sites/nhtsa.gov/files/812166-2014automaticemergencybrakingtesttrackeval.pdf

Harless, D.W. and Hoffer, G.E. (2007). Do laboratory frontal crash test programs predict driver fatality risk? Evidence from within vehicle line variation in test ratings. Accident Analysis and Prevention. 39:902–913.

Highway Loss Data Institute. (2020). HILDI automobile size and class definitions. Arlington, VA. May 2020.

HILDI (2020). Compendium of HILDI collision avoidance research. https://www.iihs.org/media/e635cc76-b9bc-4bad-a30a-5d7b78791df2/vxeQ3A/HLDI%20Research/Collisions%20avoidance%20features/37-12-compendium.pdf

Homedy, J. (2022). Submission to Docket No. NHTSA-2021-0002, May 24, 2022. https://www.ntsb.gov/news/Documents/NTSB%20comments%20on%20NHTSA%20New%20Car%20Assessment%20Program%20NPRM.pdf

IIHS. (No date). Vehicle ratings. https://www.iihs.org/?gclid=Cj0KCQjwsrWZBhC4ARIsAGGUJuqMz9wBRzEFFB9PKQeqE5h4NbkPW7QtTRvciroulTi7b_Ej9qfxXzgaAqttEALw_wcB

Insurance Institute for Highway Safety. (2020). Driver death rates remain high among small cars. Status Report, May 28. https://www.iihs.org/api/datastoredocument/status-report/pdf/55/2

Insurance Institute for Highway Safety. (No date). 2022 Top Safety Picks. https://www.iihs.org/ratings/top-safety-picks#criteria

Insurance Institute for Highway Safety. (No date). About our tests. https://www.iihs.org/ratings/about-our-tests

Insurance Institute for Highway Safety. (No date). Vehicle size and weight. https://www.iihs.org/topics/vehicle-size-and-weight

Lefler, D.E. and Gabler, H.C. (2004). The fatality and injury risk of light truck impacts with pedestrians in the United States. Accident Analysis and Prevention, 36:295–304. https://grist.org/wp-content/uploads/2005/10/aap-2004.pdf

Lidbe, A. et al. (2020). Do NHTSA vehicle safety ratings affect side impact crash outcomes? Journal of Safety Research 73:1–7. https://www.sciencedirect.com/science/article/abs/pii/S0022437520300074?via%3Dihub

Mittlbock, M. and Heinzl, H. (2001) A note on R^2^ measures for Poisson and logistic regression models when both models are applicable. Journal of Clinical Epidemiology 54:99–103. https://cemsiis.meduniwien.ac.at/fileadmin/user_upload/_imported/fileadmin/msi_akim/CeMSIIS/KB/volltexte/Mittlboeck_Heinzl_2001_Journal_of_Clinical_Epidemiology.pdf

Monfort, S.S. and Nolan, J.M. (2019) Trends in aggressivity and driver risk for cars, SUVs, and pickups: Vehicle incompatibility from 1989 to 2016. Traffic Injury Prevention, 20:sup1, S92–S96, DOI: 10.1080/15389588.2019.1632442

Mueller, A.S., Reagan, I.J. and Cicchino, J.B. (2021). Addressing Driver Disengagement and Proper System Use: Human Factors Recommendations for Level 2 Driving Automation Design. Journal of Cognitive Engineering and Decision Making. https://journals.sagepub.com/doi/full/10.1177/1555343420983126

National Highway Traffic Safety Administration. (2019). NHTSA announces updates to historic AEB commitment by 20 automakers. U.S. Department of Transportation. December 17. NHTSA Announces Update to Historic AEB Commitment by 20 Automakers | NHTSA

NHTSA. (No date). Research Vehicle Safety Ratings. https://www.nhtsa.gov/ratings?cmpid=TSGSNF0417&gclid=Cj0KCQjwsrWZBhC4ARIsAGGUJur5ELq6hB1_ZEvrCXW3Kuw4dCjPFq0EAjtyyJAZIrWB1trAva5P5UQaAhqUEALw_wcB&gclsrc=aw.ds

NHTSA File Downloads. (1975-2020). National Highway Traffic Safety Administration. https://www.nhtsa.gov/file-downloads?p=nhtsa/downloads/FARS/

O’Neill B., Joksch H., Haddon W. Jr. (1974). Empirical relationships between car size, car weight and crash injuries in car-to-car crashes. In: Proceedings of the Third International Congress on Automotive Safety. San Francisco, CA. Available upon request at https://www.iihs.org/topics/bibliography

Phillips, C.R. et al. (2021). An analysis of crash-safety ratings and the true assessment of injuries by vehicle. Computational Statistics. 36:1638–1660. An analysis of crash-safety ratings and the true assessment of injuries by vehicle | SpringerLink

Regan I.J. et al. (2021). Disengagement from driving when using automation during a 4-week field trial. Transportation Research Part F: Psychology and Behavior. 82:400–411. https://www.sciencedirect.com/science/article/abs/pii/S136984782100214X

Rizzi M., Kullgren A. and Tingvall C. (2014). Injury crash reduction of low□speed Autonomous Emergency Braking (AEB) on passenger cars. In: Proceedings of the 2014 International Research Council on Biomechanics of Injury (IRCOBI) Conference. Zurich, Switzerland: International Research Council on the Biomechanics of Injury. https://researchmgt.monash.edu/ws/portalfiles/portal/264082835/178609265_oa.pdf

Robertson L.S. (2006). Blood and oil: Vehicle characteristics in relation to fatality risk and fuel economy. American Journal of Public Health 96:1906–1909. https://ajph.aphapublications.org/doi/full/10.2105/AJPH.2005.084061

Robertson L.S. (2018). Reversal of the road death trend in the U.S. in 2015–2016: An examination of the climate and economic hypotheses. Journal of Transport & Health. 9:161–168. https://www.sciencedirect.com/science/article/abs/pii/S2214140518300021

Robertson, L.S. (2022). Roads to COVID-19 Containment and Spread. New York: Austin Macauley Publishers LLC.

Robertson, L.S. and Baker, S.B. (1975). Motor vehicle sizes in 1440 fatal crashes. Accident Analysis and Prevention. 8:167–175. Available upon request at https://www.iihs.org/topics/bibliography

Robertson L.S. and Maloney A. (1997). Motor vehicle rollover and static stability: an exposure study. American Journal of Public Health. 87:839–841. https://ajph.aphapublications.org/doi/pdf/10.2105/AJPH.87.5.839

Segui-Gomez, M. and Lopes-Valdez, F.J. (2007). An Evaluation of the Euroncap Crash Test Safety Ratings in the Real World. Annu Proc Assoc Adv Automot Med. 2007; 51: 281–298. https://www.sciencedirect.com/science/article/abs/pii/S000145750700005X

Selvin S. (1991). Statistical Analysis of Epidemiological Data. New York: Oxford University Press, 1991.

Tay R. (1998) New Vehicle Consumption and Fuel Efficiency: A Nested Logit Approach. Transportation Research Part E: Logistics and Transportation Review. 1998. 34:39–51. https://scholar.google.com/scholar?hl=en&as_sdt=0%2C3&q=New+Vehicle+Consumption+and+Fuel+Efficiency%3A+A+Nested+Logit+Approach+&btnG=

Tyndall, J. (2021) Pedestrian deaths and large vehicles. Economics of Transportation, 26–27:100219. https://doi.org/10.1016/j.ecotra.2021.100219

U.S. Department of Transportation. Annual. Highway Statistics. (1978-2021). Washington, DC. https://www.fhwa.dot.gov/policyinformation/statistics.cfm

Wang X. and Kockleman K.M. (2005). Use of heteroscedastic ordered logit model to study severity of occupant injury: Distinguishing effects of vehicle weight and type. Transportation Research Record: Journal of the Transportation Research Board 1908(1):195–204. https://journals.sagepub.com/doi/abs/10.1177/0361198105190800124

White, M. J. (2004). The “arms race” on American roads: The effect of SUV’s and pickup trucks on traffic safety. The Journal of Law and Economics. 47:333–355. (PDF) The “Arms Race” on American Roads: The Effect of Sport Utility Vehicles and Pickup Trucks on Traffic Safety (researchgate.net)

